# Dietary exposures and risk of anxiety and depression symptoms in the Lothian Birth Cohort 1936: a cohort-level GLAD Project analysis

**DOI:** 10.1101/2025.10.07.25337499

**Authors:** Janie Corley, Paul Redmond, Melissa M Lane, Felice N Jacka, Deborah N Ashtree, Simon R Cox

## Abstract

**Background:** Mental disorders such as anxiety and depression are leading contributors to disability worldwide, with growing concern in older adults—particularly in Scotland—where poor diet and mental health challenges are common. While the Global Burden of Disease (GBD) study has identified dietary risk factors for physical health, their associations with mental health in later life remain underexplored.

**Methods:** We used data from 880 participants (mean age 70y) of the Lothian Birth Cohort 1936 in Scotland to examine cross-sectional associations between 11 GBD-defined dietary exposures (fruit, vegetables, legumes, wholegrains, milk, fibre, calcium, polyunsaturated fatty acids, red meat, processed meat, sugar-sweetened beverages) and symptoms of anxiety and depression. Analyses followed harmonised protocols from the Global burden of disease Lifestyle And mental Disorders (GLAD) project (DERR2-10.2196/65576). Dietary exposures were standardised (g/day or % kcal/day), and mental health symptoms dichotomised using the Hospital Anxiety and Depression Scale (HADS ≥8 vs <8). Logistic regression models estimated odds ratios per one standard deviation increase, unadjusted and adjusted for age, sex, and education. Sensitivity analyses used energy-adjusted dietary intakes.

**Results:** Higher milk and calcium intakes were associated with greater odds of anxiety (milk adjusted OR = 1.194, 95% CI 1.018–1.400; calcium adjusted OR = 1.210, 95% CI 1.027–1.426), and higher sugar-sweetened beverage intake with greater odds of depression (adjusted OR = 1.243, 95% CI 1.007–1.535). After energy-adjustment, higher milk intake remained associated with anxiety (adjusted OR = 1.196, 95% CI = 1.020–1.403), higher fruit intake was associated with lower odds of anxiety (adjusted OR = 0.792, 95% CI 0.640–0.981), and sugar-sweetened beverages remained associated with depression (adjusted OR = 1.249, 95% CI 1.013–1.541). No other associations were significant.

**Conclusions:** In older Scottish adults, we observed only modest evidence that certain GBD-defined dietary exposures are associated with anxiety and depression symptoms.

## Introduction

Common mental disorders (CMDs), including depression and anxiety, are leading contributors to global disease burden, disability, and economic cost (GBD 2019 Mental Disorders Collaborators, 2022). CMDs account for approximately 5% of global disability-adjusted life years (DALYs), with their economic impact projected to exceed $16 trillion by 2030 (Arias et a., 2022). While established risk factors such as genetic vulnerability and early-life adversity are important considerations in the aetiology of CMDs, there is growing recognition of the role of modifiable lifestyle factors—especially diet—as promising targets for CMD prevention and treatment (Firth et al., 2020).

Emerging evidence links aspects of dietary quality to CMD risk: diets rich in fruits, vegetables, whole grains, and fibre are associated with lower risk of depression and anxiety, while high intakes of processed foods, saturated fats, and added sugars are linked to poorer mental health (Firth et al., 2020; Opie et al., 2017; Li et al., 2017; Marx et al., 2017; O’Neil et al., 2014; Saghafian et al., 2018). However, heterogeneity in study designs, dietary assessment methods, and limited representation of older adults and geographic diversity, have constrained the generalisability of findings (Marx et al., 2017).

In Scotland, poor diet quality remains a significant public health concern, particularly among older adults who experience higher rates of diet-related chronic disease and mental health issues than their peers in other UK regions (Food Standards Scotland, 2022). While this highlights an urgent need to identify dietary risk factors for CMD in later life in Scotland, it also reflects a wider gap: despite increasing evidence demonstrating associations between diet and mental health globally, dietary exposures remain underrepresented in burden estimates produced by initiatives such as the Global Burden of Disease (GBD) study (GBD 2019 Risk Factors Collaborators, 2020).

To address this gap, the Global burden of disease Lifestyle And mental Disorder (GLAD) Taskforce (Ashtree et al., 2025) was launched to quantify the contribution of lifestyle factors—including diet— to CMDs using harmonised methodologies across international cohorts. The Lothian Birth Cohort 1936 (LBC1936), a well-characterised sample of older Scottish adults with detailed mental health, dietary, and sociodemographic data, is uniquely placed to contribute to this initiative. The cohort’s narrow age range and shared sociohistorical background offer valuable insights into ageing-related dietary and mental health patterns.

In this study, we examine associations between 11 GBD-defined dietary exposures and symptoms of anxiety and depression in the LBC1936. Our findings contribute cohort-level evidence to the GLAD initiative and aim to inform age- and context-appropriate dietary strategies for mental health promotion in later life.

## Methods

### Participants

The present study draws on baseline data from the Lothian Birth Cohort 1936 (LBC1936), a longitudinal study of ageing based in Scotland. The cohort comprises 1091 community-dwelling individuals (548 men and 543 women), all born in 1936, most of whom were residing in Edinburgh and the surrounding Lothian region at the time of recruitment in later life. Baseline assessment for the current study took place between 2004 and 2007, when participants were approximately 70 years old (mean age = 69.5 years, SD = 0.8). At this wave, participants completed a comprehensive battery of cognitive, biomedical, psychosocial, and lifestyle assessments, including detailed dietary and mental health questionnaires. For the current analyses, those eligible for inclusion were N = 882 participants who completed the dietary questionnaire at baseline.

Recruitment and assessment procedures for the cohort have been described in detail elsewhere (Deary et al., 2007; Taylor et al., 2018). All procedures were conducted in accordance with the ethical standards set out in the Declaration of Helsinki, and ethical approval was obtained from the Multi-Centre Research Ethics Committee for Scotland (MREC/01/0/56) and the Lothian Research Ethics Committee (LREC/2003/2/29). All participants provided written informed consent.

### Assessment of Dietary Exposures

Dietary data were collected using the Scottish Collaborative Group 168-item Food Frequency Questionnaire (FFQ), version 7.0 (Masson et al., 2003) at baseline. This semi-quantitative FFQ was specifically designed for use in older adult populations and has been validated for use in community-dwelling older individuals in Scotland (https://www.abdn.ac.uk/iahs/research-support/food-frequency/). The FFQ assesses habitual dietary intake over the previous 2–3 months by asking participants to report the typical frequency and quantity of consumption for a wide range of foods and drinks. Each item specifies a common unit or portion size, and responses are recorded using a nine-point scale ranging from “rarely or never” to “7 or more times per day.” Participants completed the FFQ at home and returned it by post. The repeatability and validity of the FFQ have been established by comparison with 4-day weighed food diaries, showing reasonable stability in short-term dietary intake and good validity for most nutrients in older populations (Masson et al., 2003; 2004; Jia et al., 2008).

We used data for the following GBD-defined dietary categories: fruit, vegetables, legumes, wholegrains, milk, fibre, calcium, polyunsaturated fatty acids (PUFAs), red meat, processed meat, sugar-sweetened beverages, according to the ‘Dietary Exposure Definitions’ in the GLAD Taskforce analysis protocol (Ashtree et al., 2025). Intakes of individual foods within each category were summed to obtain daily intake of each category, represented in grams (g) per day. Estimations of fibre (g/day), calcium (g/day), and polyunsaturated fats (PUFAs) were extracted. PUFAs were represented as a percentage of total daily energy intake (% kcal/day). Dietary exposures were operationalised in line with GBD dietary risk definitions (GBD 2019 Risk Factors Collaborators, 2020). Participants were classified as “at risk” if their intake fell below or above the following thresholds: <310–340 g/day fruit; <280–320 g/day vegetables; <90–100 g/day legumes; <140–160 g/day wholegrains; <360–500 g/day milk; <21–22 g/day fibre; <1.06–1.1 g/day calcium; <7–9% total energy from polyunsaturated fat; or any intake of red meat, processed meat, or sugar-sweetened beverages.

### Assessment of Anxiety and Depression Outcomes

Symptoms of anxiety and depression were assessed using the Hospital Anxiety and Depression Scale (HADS), a widely used 14-item self-report questionnaire designed to detect psychological distress in non-psychiatric populations (Zigmond & Snaith, 1983). The scale comprises two subscales: one for anxiety (HADS-A) and one for depression (HADS-D), each consisting of 7 items scored from 0 to 3, with total subscale scores ranging from 0 to 21. In accordance with the GLAD Taskforce protocols, a score of ≥8 on either subscale was used as the cut-off to indicate likely presence of the corresponding condition (i.e., anxiety or depression). This threshold has been shown to have good sensitivity and specificity for identifying probable cases in older adult populations; HADS-A at this cut-off demonstrates a sensitivity of approximately 0.90 and specificity of about 0.78, while HADS-D yields sensitivity around 0.83 and specificity near 0.79 (Bjelland et al., 2002). However, we refer to these outcomes as symptoms of anxiety and symptoms of depression throughout.

### Assessment of Covariates

Covariates included age in days, sex, and years of education, all of which are routinely collected and standardly coded in the LBC1936. Age was calculated precisely in days from date of birth to the date of assessment at age 70. Sex was recorded as male or female based on participant self-report at recruitment. Education was measured as the total number of years of formal full-time schooling, derived from a detailed educational history questionnaire administered at baseline. These covariates were selected based on their established relevance to both dietary behaviours and mental health outcomes in older adults, and in accordance with the GLAD Taskforce protocols for modelling.

### Statistical Analysis

These analyses were undertaken to contribute to a GLAD Taskforce meta-analysis and proceed with the specific model sets to maximise cross-cohort harmonisation for that specific purpose.

All statistical analyses were conducted in R version 4.3.3 (R Core Team, 2024). Binary logistic regression was used to examine associations between 11 dietary exposures and symptoms of anxiety and depression, analysed separately using the respective HADS subscale (HADS <8 = 0, HADS ≥8 = 1). Dietary predictors were standardised (z-scored) so that associations were reported per 1 standard deviation (SD) increase, aiding interpretability and comparability of effect sizes. Analyses were first conducted using raw (unadjusted for energy intake) values, and then repeated using energy-adjusted predictors, as sensitivity analyses. For energy adjustment, the residual method was applied as described by Willett et al. (1997). Specifically, each dietary intake variable (excluding PUFA) was regressed on total energy intake (kilocalories/day), and the resulting residuals were extracted and standardised. These standardised residuals were then used as predictors in the regression models.

Four logistic regression models were run for each dietary exposure: unadjusted and adjusted (for age, sex, and years of education), for each outcome (anxiety and depression symptoms). Age and education were standardised (z-scored) prior to modelling. Model assumptions for logistic regression were checked and upheld. Odds ratios (ORs), 95% confidence intervals (CIs), and p-values were reported. To account for multiple comparisons, we applied a False Discovery Rate (FDR; Benjamini & Hochberg, 1995) correction across all models with a significance threshold of q < 0.05.

## Results

### Participant Characteristics

A total of 882 participants completed the FFQ. Two participants without HADS data were excluded, leaving an analytic sample of N = 880 (52% women, 48% men) with a mean age of 70 years (SD = 0.8). The characteristics of the study participants are presented in Table 1. Based on the GLAD protocol cut-off scores, N = 164 (19%) had probable anxiety and N = 31 (4%) had probable depression.

**Table 1.**
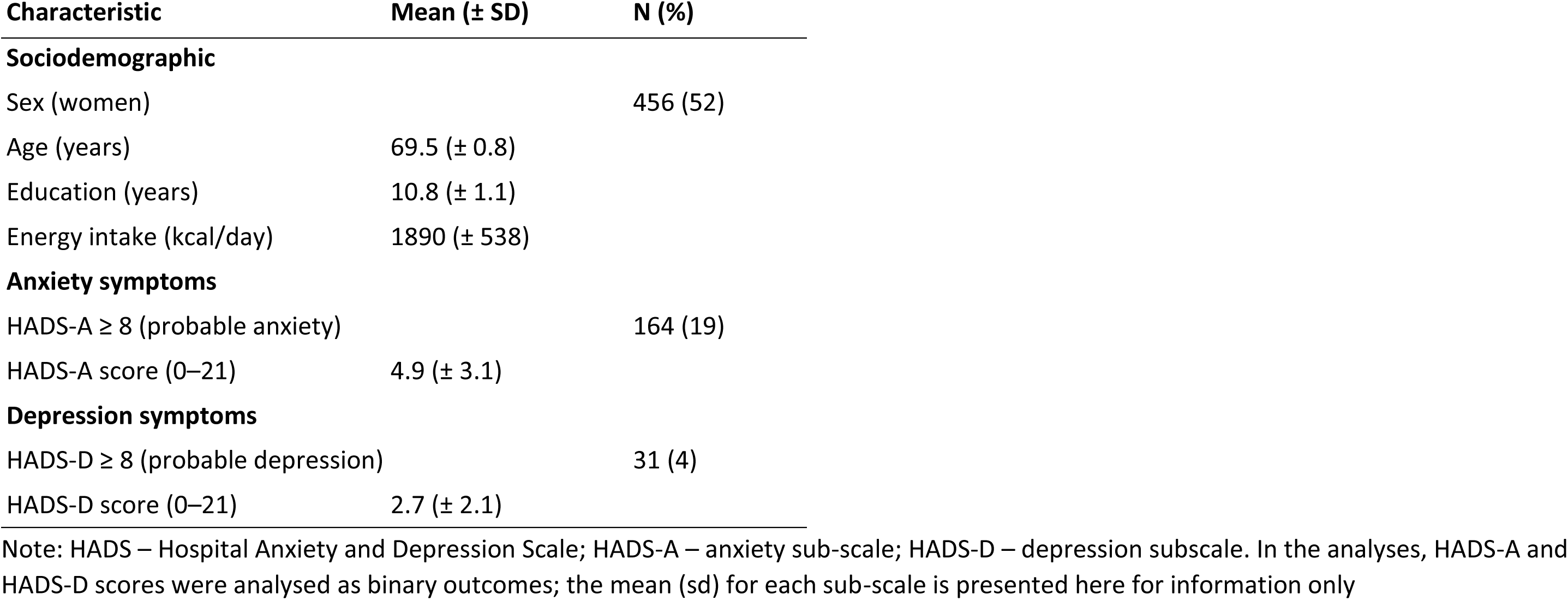
Participant characteristics (Lothian Birth Cohort 1936, N = 880)

| Characteristic | Mean (± SD) | N (%) |
| --- | --- | --- |
| **Sociodemographic** |  |  |
| Sex (women) |  | 456 (52) |
| Age (years) | 69.5 (± 0.8) | |
| Education (years) | 10.8 (± 1.1) | |
| Energy intake (kcal/day) | 1890 (± 538) | |
| **Anxiety symptoms** |  |  |
| HADS-A ≥ 8 (probable anxiety) | | 164 (19) |
| HADS-A score (0–21) | 4.9 (± 3.1) | |
| **Depression symptoms** |  |  |
| HADS-D ≥ 8 (probable depression) | | 31 (4) |
| HADS-D score (0–21) | 2.7 (± 2.1) | |

Note: HADS – Hospital Anxiety and Depression Scale; HADS-A – anxiety sub-scale; HADS-D – depression subscale. In the analyses, HADS-A and HADS-D scores were analysed as binary outcomes; the mean (sd) for each sub-scale is presented here for information only

Table 2 presents the mean daily intakes of available dietary exposures in the LBC1936 sample at mean age 70, benchmarked against GBD-defined risk thresholds. On average, participants did not meet the optimal intake levels for fruit, vegetables, legumes, wholegrains, milk, fibre, calcium, or polyunsaturated fat, with between 61.9% (calcium) and 100% (polyunsaturated fat) of participants classified as “at risk” according to GBD definitions. In addition, the sample mean exceeded the recommended thresholds for red and processed meats and sugar-sweetened beverages, where 94.5%, 94.0%, and 34.9% of participants, respectively, were at risk. These patterns, presented as a bar plot in Figure 1, suggest a broadly suboptimal dietary profile in this older adult cohort, characterised by widespread insufficient consumption of ‘healthy’ foods and excess intake of ‘unhealthy’ dietary components.

**Figure 1.**
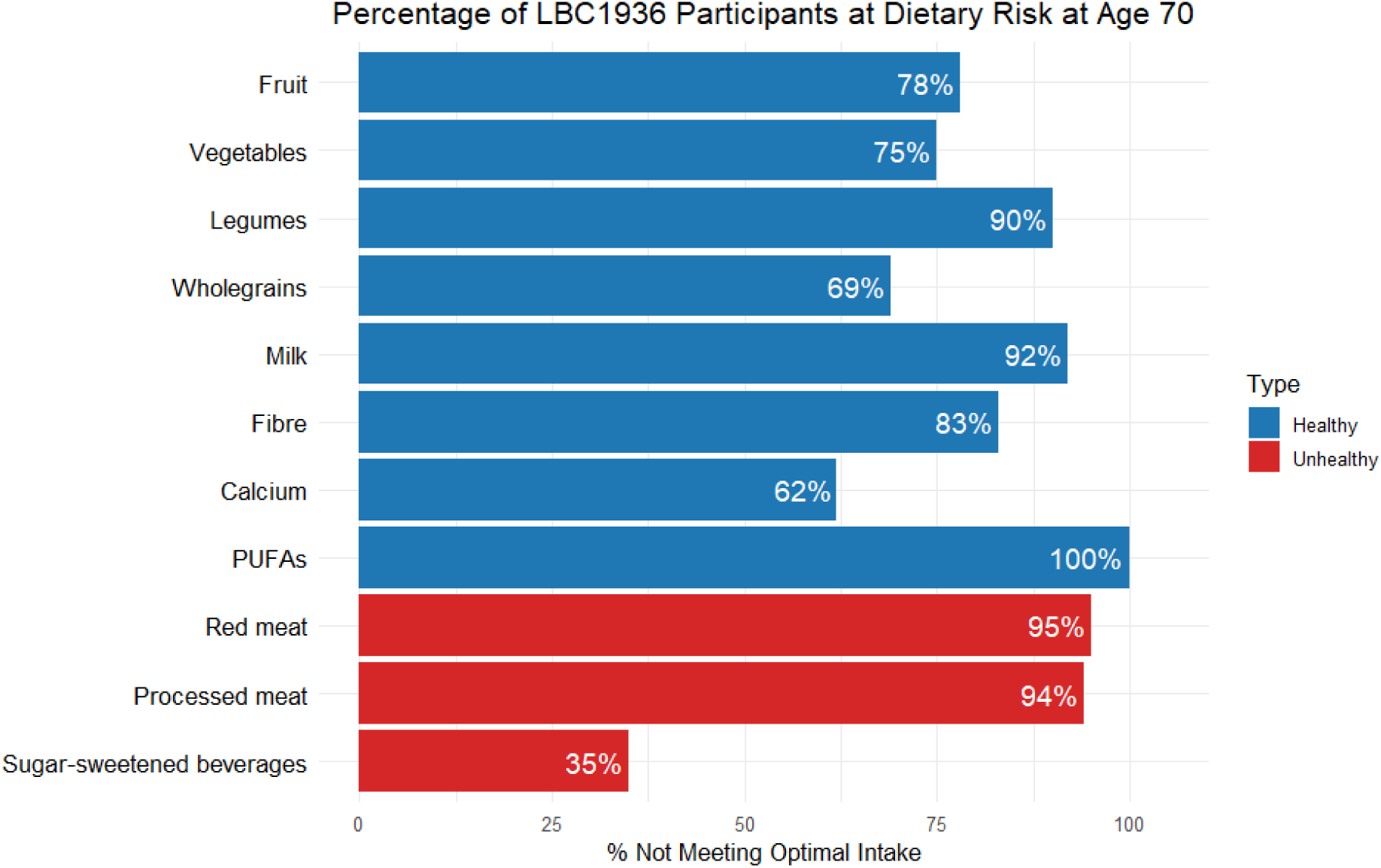
Percentage of participants at age 70 in the Lothian Birth Cohort 1936 not meeting the Global Burden of Disease (GBD) optimal intake levels for selected dietary exposures. Putatively ‘healthy’ exposures (fruit, vegetables, legumes, wholegrains, milk, fibre, calcium, PUFAs - polyunsaturated fatty acids) are shown in blue; putatively ‘unhealthy’ exposures (red meat, processed meat, sugar-sweetened beverages) are shown in red. Bars represent the proportion of the sample with suboptimal intakes; higher percentages indicate poorer dietary adherence to GBD recommendations. Values correspond to those presented in Table 2 (% at risk).

**Table 2.** Dietary exposures in the LBC1936 at age 70y, based on GBD dietary risk definitions.

| **Dietary exposure** | **Risk Factor Definition** | **Daily mean intake (± SD)** | **% At Risk** |
| --- | --- | --- | --- |
| Fruit | Diet low in fruits (<310–340 g) | 218 g (±182) | 77.5% |
| Vegetables | Diet low in vegetables (<280–320 g) | 236 g (±195) | 74.4% |
| Legumes | Diet low in legumes (<90–100 g) | 45 g (±38) | 89.8% |
| Wholegrains | Diet low in whole grains (<140–160 g) | 104 g (±125) | 69.3% |
| Milk | Diet low in milk (<360–500 g) | 195 g (±170) | 92.0% |
| Red meat | Diet high in red meat (any intake) | 47 g (±35) | 94.5% |
| Processed meat | Diet high in processed meat (any intake) | 24 g (±21) | 94.0% |
| Sugar-sweetened beverages | Diet high in sugar-sweetened beverages (any intake) | 46 g (±119) | 34.9% |
| Fibre | Diet low in fibre (<21–22 g) | 16 g (±6) | 82.7% |
| Calcium | Diet low in calcium (<1.06–1.1 g) | 1.0 g (±0.4) | 61.9% |
| Polyunsaturated fat | Diet low in polyunsaturated fatty acids (<7–9% total energy intake) | 0.14% (±0.03) | 100.0% |

Note: dietary risk factors definitions are based on the GBD dietary risk exposure definitions (GBD 2019 Risk Factors Collaborators. (2020), Supplementary Material 1, page 217-218). % At Risk reflects the % of the overall sample with suboptimal dietary intakes (GBD definition).

### Diet and Anxiety Symptoms

Higher milk intake was positively associated with anxiety symptoms. In unadjusted models, each standard deviation (SD) increase in milk intake was associated with higher odds of anxiety symptoms before (unadjusted OR = 1.182, 95% CI = 1.012–1.380, *p* = 0.035) and after adjusting for age, sex, and years of education (adjusted OR = 1.194, 95% CI = 1.018–1.400, *p* = 0.029). Similarly, higher calcium intake was positively associated with anxiety symptoms. In unadjusted models, each SD increase in calcium intake was associated with higher odds of anxiety symptoms (unadjusted OR = 1.189, 95% CI = 1.013–1.396, *p* = 0.034), and this association persisted in adjusted models (adjusted OR = 1.210, 95% CI = 1.027–1.426, *p* = 0.023). Results for the original (non-energy-adjusted) dietary exposures are presented in Table S1.

In sensitivity analyses, energy-adjusted milk intake was also associated with higher odds of anxiety before and after covariate adjustment (unadjusted OR = 1.185, 95% CI = 1.015–1.384, p = 0.032; adjusted OR = 1.196, 95% CI = 1.020–1.403, *p* = 0.028), while energy-adjusted fruit intake was associated with lower odds before and after covariate adjustment (unadjusted OR = 0.798, 95% CI = 0.648, 0.982, *p* = 0.033; adjusted OR = 0.792, 95% CI = 0.640–0.981, *p* = 0.032). The positive association between energy-adjusted calcium intake and anxiety symptoms observed in unadjusted models (OR = 1.194, 95% CI = 1.018–1.400, *p* = 0.029) was attenuated following covariate adjustment (OR = 1.128, 95% CI = 0.952, 1.335, *p* = 0.163) and no longer statistically significant. Results for the energy-adjusted dietary exposures are shown in Table S2.

None of the other dietary exposures, including vegetables, legumes, wholegrains, sugar-sweetened beverages, fibre, or PUFAs, were associated with anxiety symptoms in any model.

### Diet and Depression Symptoms

Intake of sugar-sweetened beverages was consistently associated with depressive symptoms. In unadjusted models, each SD increase in sugar-sweetened beverage consumption was associated with higher odds of depressive symptoms before (unadjusted OR = 1.259, 95% CI = 1.023–1.549, *p* = 0.029) and after adjusting for covariates (adjusted OR = 1.243, 95% CI = 1.007–1.535, *p* = 0.043). Results for the original (non-energy-adjusted) dietary exposures are presented in Table S3. Findings were similar when sugar-sweetened beverage intake was energy-adjusted. Energy-adjusted sugar-sweetened beverage intake was associated with higher odds of depressive symptoms in both unadjusted (OR = 1.262, 95% CI = 1.026–1.553, *p* = 0.027) and adjusted models (adjusted OR = 1.249, 95% CI = 1.013–1.541, *p* = 0.038). Results for the energy-adjusted dietary exposures are presented in Table S4. No other dietary exposures were associated with depressive symptoms in any model.

Overall, none of the associations between diet and mental health (i.e., across anxiety or depression models) survived correction for multiple comparisons. Figure 2 presents the odds ratios and 95% confidence intervals for all tested associations in forest plot format.

**Figure 2.**
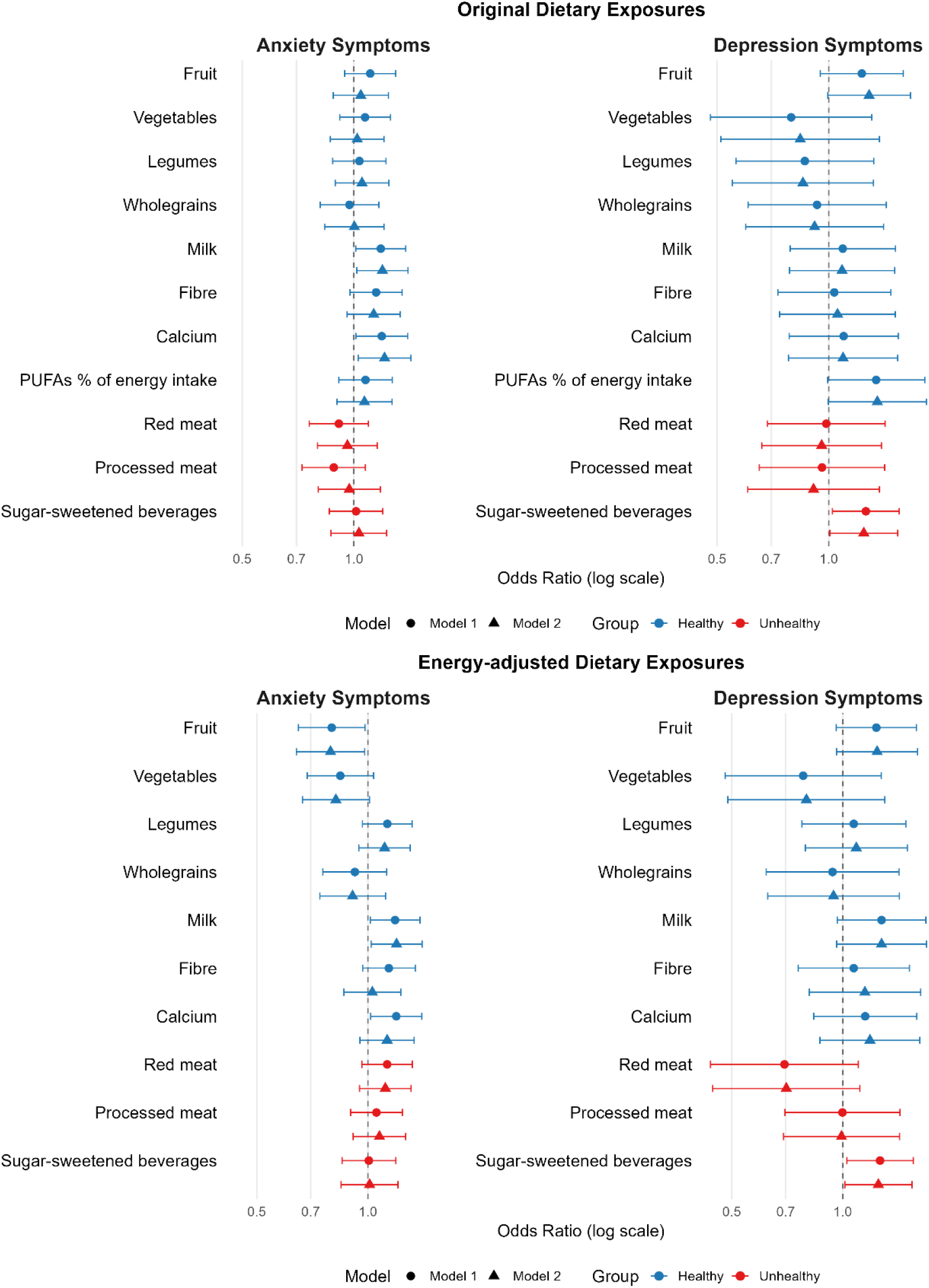
Forest plots of associations between dietary exposures and symptoms of anxiety and depression. Odds ratios (ORs) with 95% confidence intervals (CIs) are displayed on a logarithmic axis for symmetrical presentation. Two models are shown per exposure: Model 1 (unadjusted, top) and Model 2 (adjusted for age, sex, and education, below). The vertical dashed line at OR = 1.0 indicates no association; CIs crossing this line denote non-significant results. ORs >1 indicate higher odds of symptoms; ORs <1 indicate lower odds. Blue points represent putatively ‘healthy’ exposures; red points represent putatively ‘unhealthy’ exposures. Separate panels show anxiety and depression outcomes.

## Discussion

This study investigated associations between 11 dietary components—defined by the GBD framework—and symptoms of anxiety and depression in a well-characterised cohort of older Scottish adults from the Lothian Birth Cohort 1936 (LBC1936). Several nominal associations emerged. Higher milk intake was consistently associated with higher odds of anxiety symptoms, including in fully adjusted and sensitivity models. Calcium intake was also associated with higher odds of anxiety symptoms, but not after full adjustment for total energy intake and sociodemographic factors. In contrast, greater energy-adjusted fruit intake was associated with lower odds of anxiety symptoms. For depressive symptoms, higher sugar-sweetened beverage consumption was consistently associated with higher odds, both before and after energy adjustment. Although effect sizes were small, point estimates were broadly consistent across models and exposures, mostly aligning with hypothesised relationships.

The observed positive association between milk intake and anxiety symptoms in this cohort of 70-year-olds may seem counterintuitive, as dairy products and calcium are often promoted for their potential calming effects through roles in neurotransmission and muscle function (Du et al., 2019; Hess et al., 2015; Wendolowicz et al., 2016). However, several factors may explain this finding in older adults. First, milk consumption in later life may be confounded by dietary patterns associated with poorer mental health. For example, it may reflect a more traditional or restricted diet lacking in variety or nutrient-dense plant-based foods, which have been linked to better mental health (Saghafian et al., 2018). In addition, lactose intolerance or subclinical digestive discomfort—both more prevalent with ageing—may provoke gastrointestinal symptoms associated with anxiety via the gut–brain axis (Carabotti et al., 2015; Mayer et al., 2014). In the Scottish context, where tea consumption is culturally embedded and often accompanied by milk, higher milk intake could also serve as a proxy for greater caffeine consumption. Caffeine’s anxiogenic effects, especially at higher doses, are well documented (Liu et al, 2024) and its impact may be amplified in older adults due to slower metabolism or increased sensitivity. Alternatively, individuals with anxiety may be more inclined to consume milk for perceived soothing properties, raising the possibility of reverse causality. Overall, the association between milk (and potentially calcium) intake and anxiety symptoms may reflect a complex interplay of dietary patterns, physiological tolerance, and psychological state in older age.

From a biological perspective, while calcium is essential for neurotransmitter release and neuromuscular stability, excess calcium intake—especially from supplements or fortified products— has been associated with disturbances in intracellular calcium homeostasis, potentially increasing neuronal excitability and stress reactivity (Ströhle et al., 2003; Bhatia et al., 2020). Although the calcium–anxiety association did not remain significant in fully adjusted models, it may still signal the need for further investigation into how calcium interacts with other dietary and physiological factors in older populations.

The inverse association between fruit intake and anxiety symptoms—observed only in the energy-adjusted model—is consistent with prior studies linking fruit-rich diets to better mental health outcomes in older adults (Aucoin et al., 2021; Głąbska et al., 2020; Saghafian et al., 2018). Fruits are key sources of antioxidants, fibre, and micronutrients such as vitamin C, folate, and polyphenols, all of which have been implicated in neuroprotection and mood regulation (O’Neil et al., 2014). That this association emerged only in the energy-adjusted model suggests it may reflect the diet *quality* rather than quantity; individuals consuming more fruit relative to their total energy intake may be following healthier dietary patterns more broadly, which are more consistently associated with improved psychological wellbeing (Adjibade et al., 2017). Energy adjustment also helps to reduce confounding by total food intake or body size, potentially revealing more specific links between dietary exposures like fruit and mental health.

The association between sugar-sweetened beverage consumption and depressive symptoms aligns with previous evidence (Hu et al., 2019) and may be especially relevant in older adults, who are more vulnerable to metabolic dysfunction (Fountain et al., 2024). Recent umbrella review evidence provides support for a causal link between sugar-sweetened beverages and depression risk (Lane et al., 2024). Mechanistically, regular intake of these beverages can exacerbate insulin resistance and glycaemic instability, both linked to inflammation and higher depression risk (Lassale et al., 2019; Lopresti et al., 2013). Unlike fructose in whole foods, free sugars in these drinks are rapidly absorbed and metabolised, contributing to distinct cardiometabolic effects that may also impact mental health. Other potential pathways include adverse impacts on gut microbiota and the displacement of nutrient-rich drinks, which may result in deficiencies in B vitamins and magnesium, both essential nutrients essential for neurotransmitter synthesis and mood regulation (Jacka et al., 2017; Nikolova et al., 2021; Rao et al., 2008). Older adults may be particularly sensitive to the neurocognitive and emotional effects of blood glucose fluctuations, which tend to be more rapid and extreme following consumption of liquid sugars compared to solid carbohydrates (Wopereis et al., 2021)

Moreover, sugar-sweetened beverage intake may reflect broader dietary or lifestyle patterns associated with poorer mental health, such as lower physical activity, or socioeconomic disadvantage. Nonetheless, the persistence of this association after adjustment for covariates suggests a potentially independent relationship between sugar-sweetened beverage consumption and depressive symptoms in later life.

Despite these associations, the overall pattern of findings indicate that individual dietary components may exert only modest effects on mental health in this sample of older adults. The effect sizes were small (following Cohen, 1988) and none of the associations survived correction for multiple comparisons. The absence of robust findings may reflect true null effects, weak effects that require greater statistical power than afforded in this setting, or methodological constraints, including the cross-sectional design, possible residual confounding, and measurement limitations of food frequency questionnaires and self-reported psychological constructs. Furthermore, the relatively low prevalence of depression in the sample (4%)—substantially lower than the national average of around 9% for 65-74 years olds in Scotland (Scottish Government, 2023)—and reliance on a brief screening tool (HADS) may have constrained the ability to detect associations. In the event that these findings mainly lack statistical power, it is worth noting that the magnitude of our associations is broadly consistent with previous diet–depression studies (Xu et al., 2022), and even small effects may be meaningful at the population level (Funder & Ozer, 2019) given the high prevalence and public health burden of depression.

Our findings should be considered in light of broader dietary trends in Scotland, where poor diet quality, particularly low fruit and vegetable consumption and high intake of sugar-sweetened products, remains a persistent concern among older adults (Food Standards Scotland, 2022). According to the 2021 Scottish Health Survey, just 24% of adults met the recommended intake of five portions of fruit and vegetables per day (Scottish Government, 2022). Compared to other UK regions, Scotland continues to report higher rates of diet-related chronic disease and mental health challenges in later life, highlighting the need for targeted prevention strategies at the individual and community level. Consistent with these national trends, dietary intake in our sample was also suboptimal when evaluated against GBD definitions, suggesting that our findings may reflect both individual-level associations and broader population-level dietary risks. While the observed associations were modest, this study nonetheless contributes by situating diet–mental health links within a Scottish context, examining a generation exposed to shared cultural, socioeconomic, and nutritional conditions that may shape both dietary habits and psychological outcomes. In doing so, it provides contextually grounded insights that can inform locally relevant, age-sensitive public health discussions.

A key strength of this study is its use of the LBC1936, which provides rare, harmonisable data on diet, mental health, and sociodemographic factors in a narrow-age cohort of older adults, thereby minimising confounding by chronological age. Because participants are almost the same age, observed differences in health or behaviour are less likely to reflect age-related variation and more likely to capture the influence of other exposures of interest. These data offer a valuable opportunity to clarify age-specific dietary risks and inform broader public health strategies for ageing populations. Moreover, the use of harmonised analytic methods aligned with the GLAD Taskforce facilitates cross-cohort comparability and contributes to global efforts to identify modifiable lifestyle-based risk factors for common mental disorders.

## Conclusions

In conclusion, while our findings suggest that specific dietary exposures—particularly milk, calcium, fruit, and sugar-sweetened beverages—may be associated with symptoms of anxiety and depression in older age, they also highlight the complexity of diet–mental health relationships. These exposures could represent promising targets for supporting mental health in later life, but larger, longitudinal, and harmonised studies are needed to establish the direction of associations and strengthen causal inference. Such evidence will be crucial for developing age-appropriate dietary guidance to promote mental wellbeing in ageing populations.

## Acknowledgements

The authors would like to thank the participants of the Lothian Birth Cohort 1936, and the research team for their work in collecting, processing and providing the data for these analyses. We thank staff at the Rowett Research Institute at the University of Aberdeen for FFQ data entry. We would like to acknowledge all GLAD project members, including the GLAD Project Team based at Deakin University, the GLAD Advisory Group, and the GLAD Working Group comprised of all member studies.

## Data availability

To access the Lothian Birth Cohort data, see https://lothian-birth-cohorts.ed.ac.uk/data-access collaboration. Repository: STROBE checklist for “Dietary exposures and risk of anxiety and depression symptoms in the Lothian Birth Cohort 1936: a cohort-level GLAD Project analysis”. Zenodo. https://doi.org/10.5281/zenodo.17048222

## CRediT statement

JC: Writing — original draft, Writing — review and editing, Formal analysis, Methodology, Visualisation, Funding acquisition. PR: Data curation, Writing — review and editing. MML: Conceptualisation, Methodology, Resources, Writing - reviewing and editing. FNJ: Conceptualisation, Methodology, Writing — reviewing and editing. DNA: Conceptualisation, Methodology, Project administration, Resources, Writing — reviewing and editing. SRC: Writing — review and editing, Project Administration, Funding acquisition.

