## Supplementary Information for "Dietary exposures and risk of anxiety and depression symptoms in the Lothian Birth Cohort 1936: a cohort-level GLAD Project analysis"

Table S1. Odds Ratios for Dietary Predictors of Anxiety (HADS-A ≥8)

Table S2. Odds Ratios for Energy-Adjusted Dietary Predictors of Anxiety (HADS-A ≥8)

Table S3. Odds Ratios for Dietary Predictors of Depression (HADS-D ≥8)

Table S4. Odds Ratios for Energy-Adjusted Dietary Predictors of Depression (HADS-D ≥8)

Table S1. Odds Ratios for Dietary Predictors of Anxiety (HADS-A ≥8)

|  | **Model 1 (unadjusted)** | | | **Model 2 (+ age, sex, education)** | | |
| --- | --- | --- | --- | --- | --- | --- |
| **Dietary exposures** | **OR** | **95% CI** | **P-value** | **OR** | **95% CI** | **P-value** |
| Fruit | 1.107 | 0.944, 1.297 | 0.211 | 1.044 | 0.880, 1.239 | 0.622 |
| Vegetables | 1.072 | 0.916, 1.256 | 0.386 | 1.021 | 0.864, 1.206 | 0.810 |
| Legumes | 1.035 | 0.877, 1.221 | 0.683 | 1.052 | 0.891, 1.243 | 0.550 |
| Wholegrains | 0.973 | 0.811, 1.168 | 0.770 | 1.003 | 0.834, 1.206 | 0.973 |
| **Milk** | 1.182 | 1.012, 1.380 | **0.035** | 1.194 | 1.018, 1.400 | **0.029** |
| Red meat | 0.911 | 0.758, 1.095 | 0.320 | 0.961 | 0.798, 1.156 | 0.671 |
| Processed meat | 0.883 | 0.725, 1.074 | 0.212 | 0.972 | 0.802, 1.179 | 0.775 |
| Sugar-sweetened beverages | 1.014 | 0.859, 1.197 | 0.869 | 1.032 | 0.868, 1.226 | 0.722 |
| Fibre | 1.148 | 0.976, 1.349 | 0.096 | 1.131 | 0.959, 1.333 | 0.144 |
| **Calcium** | 1.189 | 1.013, 1.396 | **0.034** | 1.210 | 1.027, 1.426 | **0.023** |
| PUFAs % of energy intake | 1.075 | 0.911, 1.269 | 0.391 | 1.068 | 0.900, 1.268 | 0.453 |

Note: PUFAs – polyunsaturated fatty acids. P-values in boldtype are significant at the p < 0.05 level before FDR adjustment

Table S2. Odds Ratios for Energy-Adjusted Dietary Predictors of Anxiety (HADS-A ≥8)

|  | **Model 1 (unadjusted)** | | | **Model 2 (+ age, sex, education)** | | |
| --- | --- | --- | --- | --- | --- | --- |
| **Dietary exposures** | **OR** | **95% CI** | **P-value** | **OR** | **95% CI** | **P-value** |
| Fruit | 0.798 | 0.648, 0.982 | **0.033** | 0.792 | 0.640, 0.981 | **0.032** |
| Vegetables | 0.842 | 0.685, 1.036 | 0.104 | 0.819 | 0.665, 1.010 | 0.062 |
| Legumes | 1.129 | 0.967, 1.318 | 0.124 | 1.110 | 0.945, 1.303 | 0.203 |
| Wholegrains | 0.922 | 0.755, 1.125 | 0.422 | 0.909 | 0.741, 1.117 | 0.365 |
| **Milk** | 1.185 | 1.015, 1.384 | **0.032** | 1.196 | 1.020, 1.403 | **0.028** |
| Red meat | 1.128 | 0.963, 1.320 | 0.135 | 1.115 | 0.949, 1.310 | 0.186 |
| Processed meat | 1.055 | 0.898, 1.241 | 0.513 | 1.075 | 0.912, 1.266 | 0.389 |
| Sugar-sweetened beverages | 1.006 | 0.851, 1.191 | 0.934 | 1.011 | 0.846, 1.207 | 0.908 |
| Fibre | 1.140 | 0.968, 1.344 | 0.117 | 1.028 | 0.861, 1.228 | 0.758 |
| **Calcium** | 1.194 | 1.018, 1.400 | **0.029** | 1.128 | 0.952, 1.335 | 0.163 |
| PUFAs % of energy intake | - | - | - | - | - | - |

Note: PUFAs – polyunsaturated fatty acids. P-values in boldtype are significant at the p < 0.05 level before FDR adjustment. As the variable representing PUFAs is already expressed as a % of total daily energy intake, this variable is not included in the sensitivity analysis.

Table S3. Odds Ratios for Dietary Predictors of Depression (HADS-D ≥8)

|  | **Model 1 (unadjusted)** | | | **Model 2 (+ age, sex, education)** | | |
| --- | --- | --- | --- | --- | --- | --- |
| **Dietary exposures** | **OR** | **95% CI** | **P-value** | **OR** | **95% CI** | **P-value** |
| Fruit | 1.228 | 0.949, 1.588 | 0.119 | 1.285 | 0.994, 1.661 | 0.055 |
| Vegetables | 0.792 | 0.480, 1.306 | 0.361 | 0.838 | 0.512, 1.370 | 0.480 |
| Legumes | 0.862 | 0.562, 1.322 | 0.497 | 0.852 | 0.550, 1.320 | 0.475 |
| Wholegrains | 0.930 | 0.606, 1.429 | 0.742 | 0.916 | 0.597, 1.405 | 0.689 |
| Milk | 1.091 | 0.787, 1.514 | 0.601 | 1.086 | 0.783, 1.506 | 0.622 |
| Red meat | 0.985 | 0.684, 1.419 | 0.937 | 0.957 | 0.660, 1.387 | 0.815 |
| Processed meat | 0.959 | 0.650, 1.414 | 0.831 | 0.910 | 0.605, 1.370 | 0.652 |
| **Sugar-sweetened beverages** | 1.259 | 1.023, 1.549 | **0.029** | 1.243 | 1.007, 1.535 | **0.043** |
| Fibre | 1.035 | 0.729, 1.471 | 0.846 | 1.056 | 0.737, 1.512 | 0.766 |
| Calcium | 1.097 | 0.782, 1.539 | 0.593 | 1.093 | 0.779, 1.533 | 0.608 |
| PUFAs % of energy intake | 1.342 | 0.993, 1.815 | 0.056 | 1.352 | 0.997, 1.833 | 0.053 |

Note: PUFAs – polyunsaturated fatty acids. P-values in boldtype are significant at the p < 0.05 level before FDR adjustment

Table S4. Odds Ratios for Energy-Adjusted Dietary Predictors of Depression (HADS-D ≥8)

|  | **Model 1 (unadjusted)** | | | **Model 2 (+ age, sex, education)** | | |
| --- | --- | --- | --- | --- | --- | --- |
| **Dietary exposures** | **OR** | **95% CI** | **P-value** | **OR** | **95% CI** | **P-value** |
| Fruit | 1.233 | 0.960, 1.583 | 0.101 | 1.239 | 0.962, 1.595 | 0.096 |
| Vegetables | 0.781 | 0.480, 1.271 | 0.320 | 0.797 | 0.488, 1.300 | 0.363 |
| Legumes | 1.071 | 0.775, 1.482 | 0.677 | 1.088 | 0.792, 1.496 | 0.603 |
| Wholegrains | 0.938 | 0.620, 1.420 | 0.763 | 0.944 | 0.626, 1.424 | 0.785 |
| Milk | 1.274 | 0.966, 1.680 | 0.087 | 1.274 | 0.963, 1.685 | 0.089 |
| Red meat | 0.695 | 0.438, 1.102 | 0.122 | 0.703 | 0.444, 1.113 | 0.132 |
| Processed meat | 0.998 | 0.697, 1.430 | 0.993 | 0.992 | 0.691, 1.425 | 0.967 |
| **Sugar-sweetened beverages** | 1.262 | 1.026, 1.553 | **0.027** | 1.249 | 1.013, 1.541 | **0.038** |
| Fibre | 1.071 | 0.757, 1.516 | 0.698 | 1.148 | 0.811, 1.627 | 0.437 |
| Calcium | 1.150 | 0.834, 1.587 | 0.393 | 1.185 | 0.867, 1.618 | 0.287 |
| PUFAs % of energy intake | - | - | - | - | - | - |

Note: PUFAs – polyunsaturated fatty acids. P-values in boldtype are significant at the p < 0.05 level before FDR adjustment. As the variable representing PUFAs is already expressed as a % of total daily energy intake, this variable is not included in the sensitivity analysis.
